# How soon should patients be eligible for differentiated service delivery models for antiretroviral treatment? Evidence from Zambia

**DOI:** 10.1101/2021.08.25.21262587

**Authors:** Lise Jamieson, Sydney Rosen, Bevis Phiri, Anna Grimsrud, Muya Mwansa, Hilda Shakwelele, Prudence Haimbe, Mpande M Mwenechanya, Priscilla Lumano-Mulenga, Innocent Chimboma, Brooke E Nichols

**Affiliations:** Health Economics and Epidemiology Research Office (HE^2^RO), Department of Internal Medicine, Faculty of Health Sciences, University of the Witwatersrand, Johannesburg, South Africa; Department of Medical Microbiology, Amsterdam University Medical Centre, Amsterdam, the Netherlands; Department of Global Health, Boston University School of Public Health, Boston, MA, USA; Clinton Health Access Initiative, Lusaka, Zambia; International AIDS Society, Cape Town, South Africa; Ministry of Health, Lusaka, Zambia; The Centre for Infectious Disease Research in Zambia, Lusaka, Zambia

**Keywords:** differentiated service delivery (DSD) models, HIV, antiretroviral treatment, retention in care, DSD guidelines

## Abstract

**Introduction:** Attrition from HIV treatment is high during patients’ first 6 months after antiretroviral therapy (ART) initiation and patients with less than 6 months on ART are systematically excluded from most differentiated service delivery (DSD) models, which are intended to reduce attrition. Despite eligibility criteria requiring greater than 6 months on ART, some patients enroll earlier. Using routinely-collected medical record data in Zambia, we compared loss to follow-up (LTFU) among patients enrolling in DSD models early (<6 months on ART) to LTFU among those who enrolled according to guidelines (≥6 months on ART) in order to assess whether the ART experience eligibility criterion is necessary.

**Methods:** We extracted data from electronic medical records for adults (≥15 years) who initiated ART between 01/01/2019 and 31/12/2019 and evaluated LTFU, defined as >90 days late for last scheduled medication pickup, at 18 months for “early enrollers” (DSD enrolment after <6 months on ART) and “established enrollers” (DSD enrolment after ≥6 months on ART). We used a log-binomial model to compare LTFU risk between groups, adjusting for age, sex, urban/rural status, ART refill interval and DSD model.

**Results:** For 6,340 early enrollers and 25,857 established enrollers there were no important differences between the groups in sex (61% female), age (median 37 years), or setting (65% urban). ART refill intervals were longer for established vs early enrollers (72% vs 55% were given 4–6-month refills). LTFU at 18 months was 3% (192/6,340) for early enrollers and 5% (24,646/25,857) for established enrollers. Early enrollers were 41% less likely to be LTFU than established patients (adjusted risk ratio [95% confidence interval] 0.59 [0.50-0.68]).

**Conclusions:** Patients enrolled in DSD models in Zambia with < 6 months on ART were more likely to be retained in care than patients referred after they were established on ART. A limitation of the analysis is that early enrollers may have been selected for DSD participation due to providers’ and patients’ expectations about future retention. Offering DSD model entry to at least some ART patients <6 months after ART initiation may help address high attrition during the early treatment period.

## Introduction

A critical step toward achieving universal coverage of antiretroviral therapy (ART) for HIV is to support lifelong patient retention in ART programmes. Data from sub-Saharan Africa (SSA), where some 70% of the world’s ART patients reside, continue to indicate insufficient retention on ART [1], with about a fifth of all patients lost to care five years after treatment initiation [2]. A patient’s first six months after initiation are a high risk period for attrition: a Zambian study showed rates of loss to follow-up to be four-fold higher in the first six months of ART treatment compared to the period between six months and 3.5 years thereafter [3].

Since 2016, the World Health Organization (WHO) has recommended differentiated service delivery (DSD) for HIV treatment [4]. DSD models such as facility-based individual “fast track” medication pickup and community-based ART refills can increase access and remove barriers to care by adjusting the cadre of provider, location of service delivery, frequency of interactions with the healthcare system, and/or types of services offered to support long-term retention of people established on HIV treatment [5]. A recent systematic review reporting on outcomes of patients in DSD models in SSA found that retention in care of those in DSD models was generally within 5% of that for conventional care [6]. In Zambia, several DSD models have shown to have similar rates of retention as conventional care 12 months after DSD model entry [7,8]. The INTERVAL trial, a cluster-randomized, non-inferiority trial conducted in Malawi and Zambia, found that 6-month ART dispensing was non-inferior in terms of 12-month retention, compared to standard of care [8]. DSD models have consistently been found to save substantial time and money for patients themselves, and satisfaction with the models among both providers and patients has been high [8–10].

A major limitation of DSD models to date has been eligibility criteria that limit model enrollment to patients on the standard first-line ART regimen who are “stable” or “established on treatment,” defined as having been on ART for at least 6 or 12 months and having documented viral suppression [8,11–13]. Until April 2021, the WHO’s definition of “established” included at least 12 months of ART experience; new guidelines require at least 6 months on ART for DSD model eligibility [14]. Patients who are newly initiated on ART are thus systematically excluded from stable-patient-specific DSD models and from the benefits they offer. In the previously cited INTERVAL trial in Malawi and Zambia, 10% of all patients were excluded due to having initiated ART less than 6 months prior[15]. For patients not eligible for DSD models, guidelines typically require frequent visits to the healthcare facility and medication dispensing intervals of no more than 3 months [16]. In Zambia, all care is differentiated and dependent on the needs of the patient [11], but currently there is no evidence on the outcomes of patients with <6 months ART experience who enroll into DSD models that are typically reserved for stable patients.

Despite existing guidelines limiting DSD eligibility based on time on ART, in practice patients who do not meet guideline-recommended criteria are sometimes enrolled in DSD models for stable patients, due to provider decision, error or patient request. To begin to understand how such patients who are referred early to DSD models fare when participating in DSD models designed for those established on treatment, we analyzed routinely collected medical record data from Zambia to compare rates of retention among patients enrolled into DSD models earlier than guidelines recommend with retention among those who met all eligibility criteria.

## Methods

### Study population and outcomes

Data were retrospectively extracted in October 2021 from SmartCare, Zambia’s national electronic medical record system [17]. We extracted data for patients, aged 15 years or older, reported to have initiated ART between January 2019 and December 2020 at any of 692 health facilities across all 10 provinces. Zambian policy guidelines for this period required patients to be stable on ART before they are considered for DSD enrolment, with stability defined in the 2018 consolidated ART guidelines [11,12] as on ART for at least six months.

We defined patients who enrolled into a DSD model with <6 months of ART as “early enrollers”, while a comparison group of patients who enrolled into a DSD model with ≥6 months of ART as “established enrollers”. Patients on second-line ART (defined as those dispensed protease inhibitors such as lopinavir, atazanavir or ritonavir) were excluded from this analysis, as they are already known to be at high risk of attrition [18,19]. For both early and established enrollers, we assessed loss to follow-up (LTFU) at 18 months post-ART initiation, with LTFU defined as patients who were reported as “lost to follow-up” or “inactive” in the SmartCare database between 15 and 21 months after ART initiation date. “Inactive” was defined as having missed a scheduled visit by more than 30 days. Rates of LTFU were calculated for early and established enrollers and stratified by DSD model type and ART dispensing duration. DSD models, which had multiple names in the SmartCare database, were grouped into the following categories: 1) adherence groups (community adherence groups, rural/urban adherence groups); 2) extended clinic hours (DSD models designed for clinic access before/after hours or weekends, including scholar models); 3) fast-track (procedures to accelerate dispensing at clinics); 4) home ART delivery; 5) multi-month dispensing (MMD); and 6) community pick-up point (central dispensing units, community retail pharmacies, community ART distribution points, health posts, mobile ART distribution models) (Table 1).

**Table 1.**
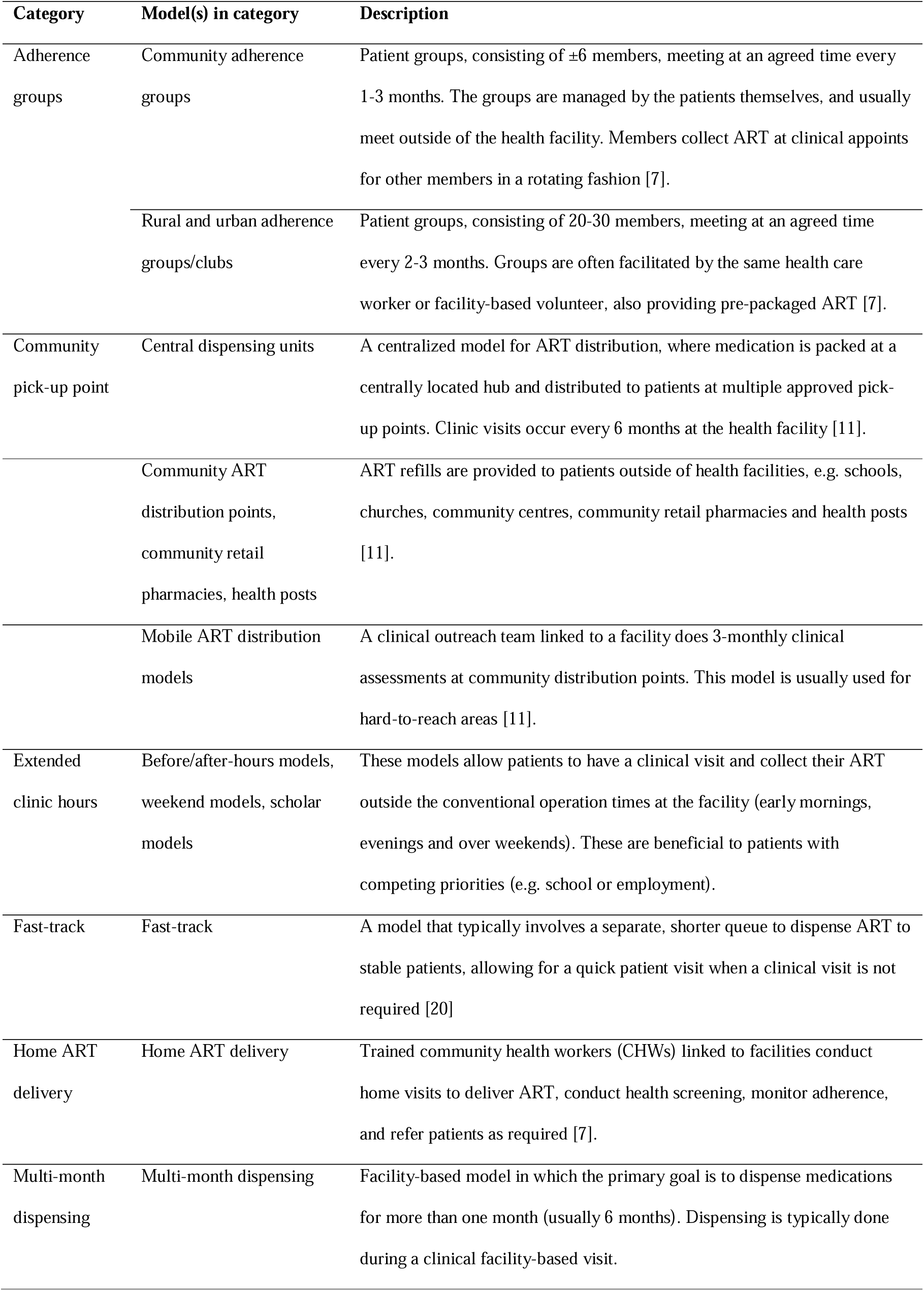
Differentiated service delivery (DSD) models for HIV treatment in use in Zambia during the study period.

| Category | Model(s) in category | Description |
| --- | --- | --- |
| Adherence groups | Community adherence groups | Patient groups, consisting of $\pm 6$ members, meeting at an agreed time every 1-3 months. The groups are managed by the patients themselves, and usually meet outside of the health facility. Members collect ART at clinical appointments for other members in a rotating fashion [7]. |
|  | Rural and urban adherence groups/clubs | Patient groups, consisting of 20-30 members, meeting at an agreed time every 2-3 months. Groups are often facilitated by the same health care worker or facility-based volunteer, also providing pre-packaged ART [7]. |
| Community pick-up point | Central dispensing units | A centralized model for ART distribution, where medication is packed at a centrally located hub and distributed to patients at multiple approved pick-up points. Clinic visits occur every 6 months at the health facility [11]. |
|  | Community ART distribution points, community retail pharmacies, health posts | ART refills are provided to patients outside of health facilities, e.g. schools, churches, community centres, community retail pharmacies and health posts [11]. |
|  | Mobile ART distribution models | A clinical outreach team linked to a facility does 3-monthly clinical assessments at community distribution points. This model is usually used for hard-to-reach areas [11]. |
| Extended clinic hours | Before/after-hours models, weekend models, scholar models | These models allow patients to have a clinical visit and collect their ART outside the conventional operation times at the facility (early mornings, evenings and over weekends). These are beneficial to patients with competing priorities (e.g. school or employment). |
| Fast-track | Fast-track | A model that typically involves a separate, shorter queue to dispense ART to stable patients, allowing for a quick patient visit when a clinical visit is not required [20] |
| Home ART delivery | Home ART delivery | Trained community health workers (CHWs) linked to facilities conduct home visits to deliver ART, conduct health screening, monitor adherence, and refer patients as required [7]. |
| Multi-month dispensing | Multi-month dispensing | Facility-based model in which the primary goal is to dispense medications for more than one month (usually 6 months). Dispensing is typically done during a clinical facility-based visit. |

### Statistical analysis

We described the demographics of our study population using descriptive statistics. We compared loss to follow-up risk between early enrollers and established enrollers and Wilson’s score interval was used to calculate 95% confidence intervals around proportions. We used a log-binomial regression to calculate risk ratios for loss to follow-up, adjusting for age, sex, urban/rural status, DSD model type and ART dispensing duration. We also conducted an age-stratified analysis and a sub-analysis restricted to facilities with a higher proportion of early enrollers, with results shown in the Supplementary Appendix.

### Ethics

This study protocol was approved by ERES Converge IRB (Zambia), protocol number 2019-Sep-030, the Human Research Ethics Committee (Medical) of the University of Witwatersrand, protocol number M190453, and the Boston University IRB H-38823 for the use of data with a waiver of consent.

## Results

### Study populations

The full SmartCare data set included 1,520,125 unique patients on ART over 2018-2021, of which 32,197 patients had enrolled into a DSD model after ART initiation and had an 18-month outcome reported within the 15-to-21-month window (Figure 1). Of these, 6,340 patients were reported to have been enrolled in DSD models <6 months after ART initiation during the study period (early enrollers). The remaining 25,857 patients comprised the comparison group of established enrollers. For early enrollers, median time enrolled in a DSD model at the time of outcome evaluation was 14.7 months (IQR 13.0-16.5); majority (81%, n=20,856) of established enrollers were on DSD models at outcome evaluation at a median of 5.8 months (interquartile range (IQR) 2.9-8.9) (Table 2). Early enrollers and established enrollers were similar with respect to age, sex and urban/rural location. Across both groups, the median age was 37 years (IQR 29 – 44), a majority (61%, 19,580/32,197) were female and most patients resided in urban settings (64%, n=20,618).

**Table 2.** Demographics of patients enrolled in differentiated service delivery models.

| Variable |  | Early enrollers<br>of DSD models<br>(N=6,340) | Established enrollers<br>of DSD models<br>(N=25,857) |
| --- | --- | --- | --- |
| **Age in years, median (IQR)** |  | 36 (29-44) | 37 (29-44) |
| **Age group** | 15-24 | 727 (11%) | 2,589 (10%) |
|  | 25-34 | 2,069 (33%) | 8,346 (32%) |
|  | 35-49 | 2,658 (42%) | 11,424 (44%) |
|  | 50+ | 885 (14%) | 3,487 (13%) |
| **Sex** | Female | 3,914 (62%) | 15,666 (61%) |
|  | Male | 2,426 (38%) | 10,191 (39%) |
| **Location** | Rural | 2,501 (39%) | 9,078 (35%) |
|  | Urban | 3,839 (61%) | 16,779 (65%) |
| **Year of ART initiation** | 2019 | 2,897 (46%) | 17,346 (67%) |
|  | 2020 | 3,443 (54%) | 8,511 (33%) |
| **DSD type** | Adherence groups | 149 (2%) | 508 (2%) |
|  | Community pickup points | 671 (11%) | 1,461 (6%) |
|  | Extended clinic hours | 85 (1%) | 97 (<1%) |
|  | Fast-track | 979 (15%) | 6,266 (24%) |
|  | Home ART delivery | 355 (6%) | 973 (4%) |
|  | Multi-month dispensing | 4,101 (65%) | 16,552 (64%) |
| **ART months dispensed** | <2 months | 636 (10%) | 1,476 (6%) |
|  | 3 months | 2,197 (35%) | 5,688 (22%) |
|  | 4-6 months | 3,507 (55%) | 18,679 (72%) |
| **Outcome Year** | 2020 | 2,863 (45%) | 17,283 (67%) |
|  | 2021 | 3,477 (55%) | 8,574 (33%) |
| **Months on ART at outcome, median (IQR)** |  | 17.9 (16.4-19.5) | 18.4 (16.7-19.8) |
| **On DSD at outcome** | Yes | 6,340 (100%) | 20,856 (81%) |
|  | No | 0 (0%) | 5,001 (19%) |
| **Months on DSD at outcome, median (IQR)** |  | 14.7 (13.0-16.5) | 5.8 (2.9-8.9) |
| **Patient outcomes by 18 months after ART initiation** | On treatment | 6,133 (97%) | 24,646 (95%) |
|  | Died | 11 (<1%) | 31 (<1%) |
|  | Lost to follow-up | 192 (3%) | 1,169 (5%) |
|  | Stopped ART | 4 (<1%) | 10 (<1%) |
|  | Stopped DSD | 0 (0%) | 1 (<1%) |

**Figure 1:**
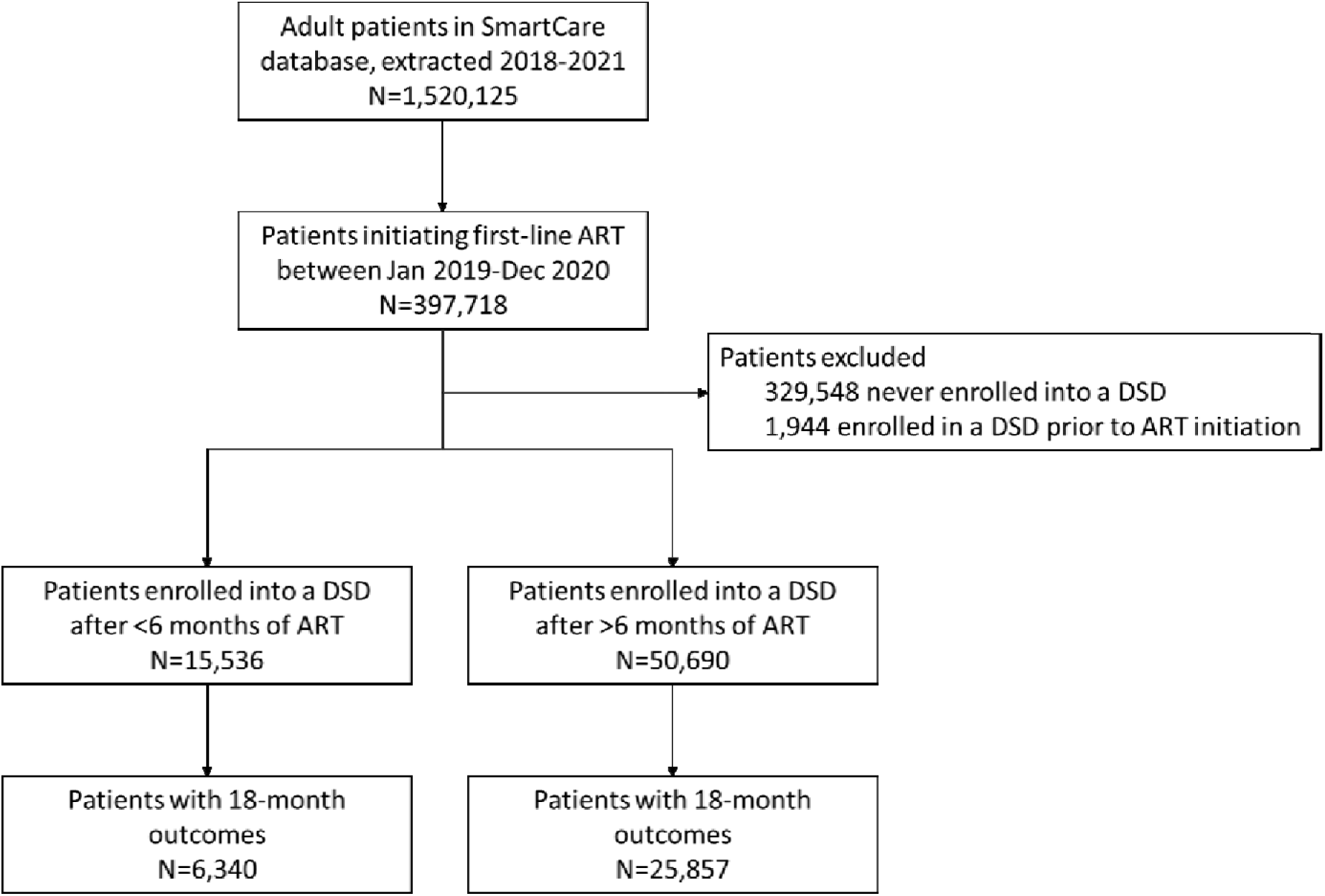
Flow diagram depicting study population.

Most patients were enrolled in either multi-month dispensing DSD models (65% [n=4,101] of early enrollers and 64% [n=16,552] of established enrollers) or fast-track (15% [n=979] of early enrollers and 24% [n=6,266] of established enrollers) (Table 1). Amongst early enrollers, around half (55%, n=3,477) were dispensed 4-6 months of ART at their most recent ART pickup, 35% (n=2,197) were dispensed 3 months of ART, and 10% (n=636) were dispensed <2 months of ART. Established enrollers had slightly longer dispensing intervals with 72% (n=18,679) dispensed 4-6 months of ART, 22% (n=5,688) dispensed 3 months of ART, and 6% (n=1,476) dispensed <2 months of ART (Table 1).

### Outcomes

Early enrollers had a slightly lower rate of loss to follow-up (3.0% [95% confidence interval (CI) 2.6%-3.5%]) compared to the established enrollers (4.5% [4.3%-4.8%]) (Table 3). Early enrollers experienced similar or lower loss to follow-up rates than established enrollers across nearly all differentiated models of care. The exception was extended clinic hours: early enrollers enrolled in the extended clinic hours model had a similar rate of loss to follow-up than established enrollers (10.6%; [5.7%-18.9%] vs. 8.2% [4.2%-15.4%], respectively). Across both early and established enrollers, longer dispensing periods were associated with lower rates of loss to follow-up, which increased from 2.5%-3.8% for 4-6-month dispensing to 3.5%-5.3% for 3-month dispensing to 4.1%-10.6% for <2-month dispensing (Table 3). Early enrollers with <2 months dispensing had a lower rate of loss to follow-up than did established enrollers (4.1%; [2.8%-5.9%] vs. 10.6% [9.1%-12.2%]).

**Table 3.** **Proportion of patients lost to follow-up at 18 months after ART initiation, stratified by differentiated service delivery (DSD) model and ART months dispensed** *(95% confidence interval in round brackets, sample numbers in square brackets)*

|  | Early enrollers | Established enrollers |
| --- | --- | --- |
| **Overall** | 3.0% (2.6% - 3.5%)<br>[192/6,340] | 4.5% (4.3% - 4.8%)<br>[1,169/25,857] |
| **By DSD model** |  |  |
| Adherence groups | 2.7% (1% - 6.7%)<br>[4/149] | 3.1% (1.9% - 5.1%)<br>[16/508] |
| Community pickup points | 4.5% (3.1% - 6.3%)<br>[30/671] | 3.3% (2.5% - 4.3%)<br>[48/1,461] |
| Extended clinic hours | 10.6% (5.7% - 18.9%)<br>[9/85] | 8.2% (4.2% - 15.4%)<br>[8/97] |
| Fast track | 3.4% (2.4% - 4.7%)<br>[33/979] | 3.6% (3.2% - 4.1%)<br>[227/6,266] |
| Home ART delivery | 1.4% (0.6% - 3.3%)<br>[5/355] | 6.3% (4.9% - 8%)<br>[61/973] |
| Multi-month dispensing | 2.7% (2.3% - 3.2%)<br>[111/4,101] | 4.9% (4.6% - 5.2%)<br>[809/16,552] |
| **By months ART dispensed** |  |  |

|  | **Early enrollers** | **Established enrollers** |
| --- | --- | --- |
| *<2 months* | 4.1% (2.8% - 5.9%)<br>[26/636] | 10.6% (9.1% - 12.2%)<br>[156/1,476] |
| *3 months* | 3.5% (2.8% - 4.4%)<br>[77/2,197] | 5.3% (4.8% - 5.9%)<br>[303/5,688] |
| *4-6 months* | 2.5% (2.1% - 3.1%)<br>[89/3,507] | 3.8% (3.5% - 4.1%)<br>[709/18,679] |

In an analysis adjusting for age, sex, location, and ART dispensing duration or DSD model type (where applicable),, early enrollers in all DSD model types and dispensing durations were 41% less likely to be lost to follow-up than established enrollers (adjusted risk ratio (aRR) 0.59 [0.50-0.68]) (Figure 2). The reduced adjusted risk of being lost to follow-up were similar for patients in adherence groups (aRR 0.79 [0.23-2.12]), multi-month dispensing (aRR 0.51 [0.41-0.61]), home ART delivery (aRR 0.18 [0.06-0.41]) and fast track models (aRR 0.74 [0.50-1.05]). Early enrollers had a statistically insignificant increased risk of being lost to follow-up in the community pick-up point (aRR 1.30 [0.81-2.03]) and extended clinic hours models (aRR 1.19 [0.43-3.34]) compared to the established enrollers.

**Figure 2.**
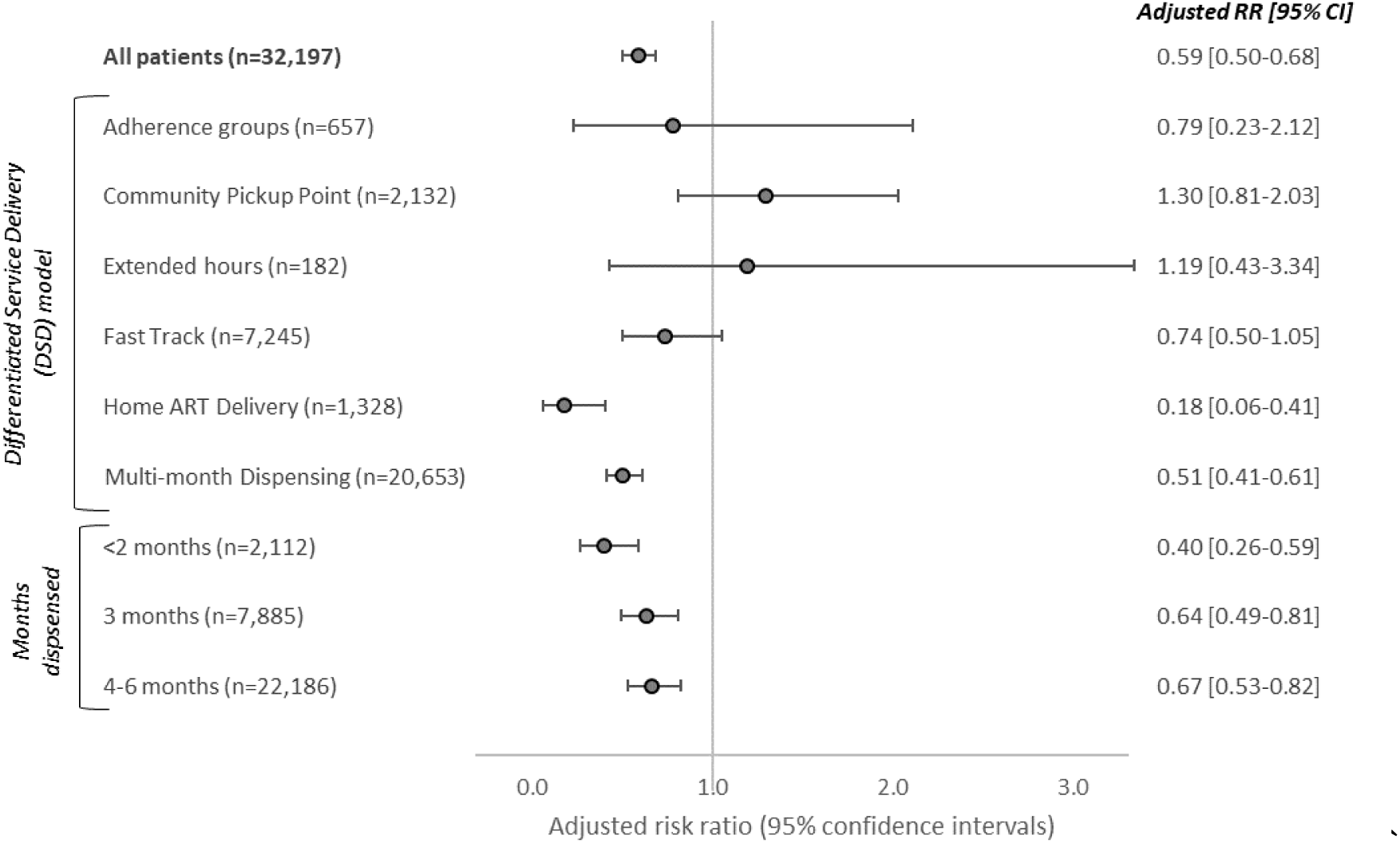
Relative risk of loss to follow-up at 18 months post-ART initiation for early enrollers of differentiated service delivery (DSD) models. (reference group: established enrollers of DSD models with >6 months of ART at DSD entry; analysis adjusted for age, sex, urban/rural status and number of months dispensed (for DSD stratified analysis) and DSD model (for months dispensed stratified analysis))

An age-stratified analysis produced similar results to the main analysis, with early enrollers in each age group being less likely to be lost to follow-up than established enrollers in the same age group. However, the effect of earlier enrollment in DSD on reduced loss to follow-up appeared less pronounced in patients on 4-6 months’ ART dispensing for those aged 25 to 49 years (Appendix Figure S1). In facilities where a larger proportion of all DSD patients enrolled in DSD models early, the trend towards early enrollers performing better persisted with respect to loss to follow-up compared to outcomes for established enrollers (Appendix Figure S2).

## Discussion

In nearly all of sub-Saharan Africa, DSD model eligibility criteria require that patients be on ART for a minimum of six months (and in some countries a minimum of 12 months) prior to DSD model enrollment [21]. We present novel data from Zambia highlighting good outcomes when newly initiated ART patients (those with less than 6 months’ ART experience) are referred early to DSD models. Those referred early to DSD appear to have good outcomes across different DSD models and age categories.

Our data begin to fill in a gap in the evidence base on the validity of time on treatment as an eligibility criterion for DSD models. Because few if any countries permit DSD model enrollment for new initiators, little evidence on their experience in DSD models has been available until now. To date, most reports on DSD outcomes have been limited to people who have spent a significant amount of time on ART prior to DSD model enrollment. In the previously mentioned INTERVAL trial, for example, participants had been on ART for a median of roughly five years at DSD model entry, while patients in a trial of multi-month dispensing in adherence clubs in South Africa had a median duration on ART of 7.3 years at baseline [22].

While ART patients in Zambia have historically been lost to follow-up at high rates in the first few months after ART initiation [3], in our DSD patient population this was less likely to be the case. Our results provide evidence to support the recent revision of WHO guidelines that reduce time on ART from 12 to six months on treatment as part the definition of “established” on ART [14]. These findings offer reassurance and evidence to countries that have expanded eligibility as they scale up DSD models [21,23], particularly to support uninterrupted access to HIV treatment during the COVID-19 pandemic, that earlier referral to DSD is possible without compromising patient care. Even if many, or most, of the patients in our “early enrollment” sample were selected deliberately because they were considered at low loss to follow-up risk, our results demonstrate that early eligibility for DSD models should be considered for at least some patients before they reach six months on ART.

Loss to follow up at 18 months after ART initiation for early and established enrollers averaged 1-11% for all six categories of DSD models studied. We did not observe any programmatically important differences by model or ART experience prior to model enrollment. Where a programmatically important difference did arise, in contrast, was in dispensing intervals. Regardless of how long a patient had been on ART at DSD model enrollment, patients who received ≤2 months of medications at a time were more likely to be lost to follow up than patients who received either 3 months or 4-6 months of medications. This likely reflects providers’ assessments of patients’ ability to remain on treatment and/or clinical condition. Those regarded as being at higher risk of attrition are asked to come to the clinic for medication refills more often, so that they can be monitored and supported more closely. Ironically, difficulty in accessing the clinic may be the very reason that some patients are at high risk of attrition. For these patients, insisting on shorter refill durations may simply exacerbate whatever challenges they face.

There were several limitations to our analysis. First, as noted above, we assume that patients with <6 months on ART in our sample were not offered DSD model enrollment at random. If providers made accurate clinical decisions about individual patients’ risks of attrition, patients in our “early enrollment” cohorts could over-represent patients thought to have low attrition risk. To achieve the results we found, providers would have had to make these decisions correctly at multiple sites across the entire country. If this is the case, our data suggest that the healthcare workers responsible for enrolling patients into DSD models can successfully identify those who will do well with early enrollment. At the same time, if the early enrollers in our data set do comprise patients at lower risk of loss to follow-up, then our results likely underestimate the true rate of loss to follow-up that would occur if early DSD enrollment were to be broadly available, without the benefit of provider selection.

A second limitation is that our data set included only patients reported in the electronic medical record system to have enrolled in a DSD model. It is possible that some patients not in DSD models may be recorded as enrolled, and some who were enrolled may have been missed. Third, bias could occur if facilities with better-than-average retention in care were also more likely to allow early DSD model enrollment. In this case, our results may reflect differences in facility quality, as well as enrollment timing. An analysis restricted to facilities with >20% early DSD enrolment showed an even lower risk of loss to follow-up among patients enrolled early into DSD models, however, compared to patients with >6 months of ART at DSD entry.

Despite these limitations, our analysis demonstrates that patients on ART for less than six months who are enrolled in existing DSD models can be successfully retained in care and may even fare better than those left in conventional care and only initiate DSD models greater than six months after ART initiation. It is likely that not all patients are ready for less intensive DSD models in their first half-year or year on treatment, but some clearly are. Since DSD models have been shown to be beneficial to patients and in some cases to providers, offering enrollment to newly-initiating ART patients may improve ART programs in general. Future research should look more closely at which patients can be enrolled early and which models of care serve these patients best.

## Conclusions

Current policy for DSD model eligibility criteria in Zambia, as in other countries, have required a minimum of 12 months of ART before a patient is considered for DSD enrolment, and more recently, a minimum of six months of ART. In order to change guidelines to allow DSD enrolment sooner after ART initiation (i.e., 6 months or less), large-scale observational evidence, implementation research or trial data demonstrating good patient outcomes among those who enrol in DSD models < six months’ post ART initiation would be required. This analysis therefore provides a critical first step towards the reassessment of the delayed DSD enrolment policies, and signals that further research needs to be conducted in other SSA countries to evaluate patient outcomes for early DSD model enrolment.

## Data Availability

Raw data were obtained from Zambia's national electronic medical record system. Derived data supporting the findings of this study are available from the corresponding author [BEN] on request.

## Competing interests

We declare no competing interests.

## Authors’ contributions

LJ, BN, SR and AG conceptualized the study. BP, HS, PH, MM, PLM, IC curated data for the study. BP, HS, PH, MMM provided supervision of the study. LJ led data analysis and drafted the paper along with BN, SR and AG. All authors contributed to data interpretation and critically reviewed a revised draft of the manuscript. All authors have read and approved the final manuscript.

## Funding

Funding for the study was provided by the Bill & Melinda Gates Foundation through OPP1192640 to Boston University. The funder had no role in study design, data collection and analysis, decision to publish or preparation of the manuscript.

## Data sharing

The data is owned by the Zambian Ministry of Health and the use of it was approved by the Human Research Ethics Committee (University of Witwatersrand, Johannesburg, South Africa) and ERES Converge IRB (Zambia). All relevant data is included in the paper and supplementary tables. The full data are available upon approval from Zambian Ministry of Health and appropriate ethics committees.

## Notes

### Competing Interest Statement

The authors have declared no competing interest.

### Summary of Updates

Re-analysis on a more recent, fuller dataset from routinely-collected electronic medical record data from Zambia.

