## Supplementary Appendix for "How soon should patients be eligible for differentiated service delivery models for antiretroviral treatment? Evidence from Zambia"

**Figure S1. Relative risk of loss to follow-up within 18 months of ART initiation for early enrollers of DSD models (ie. after <6 months of ART), stratified by dispensing period and age group** (reference group: established enrollers of DSD models with >6 months of ART at DSD enrolment; analysis adjusted for sex and urban/rural status)

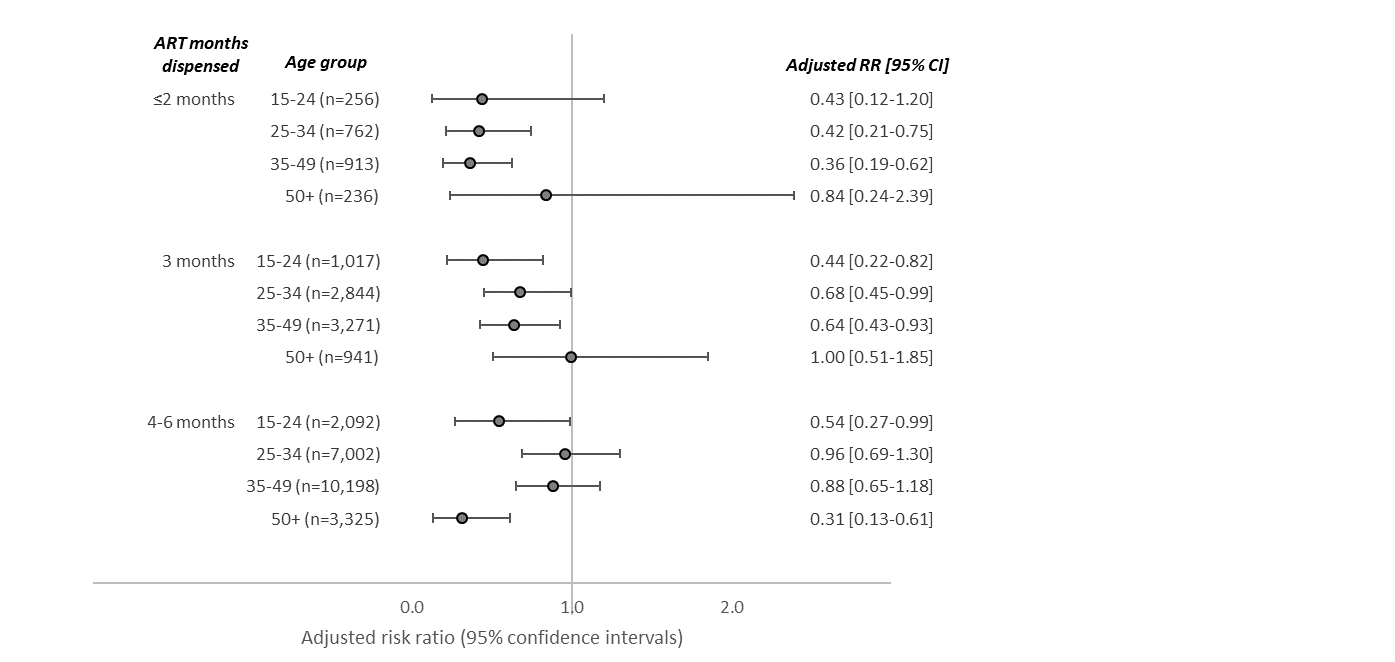

**Figure S2. Proportion loss to follow-up by time on ART at DSD entry, limited to N=37 facilities with >20% of DSD patients at each facility classified as “early enrollers”**

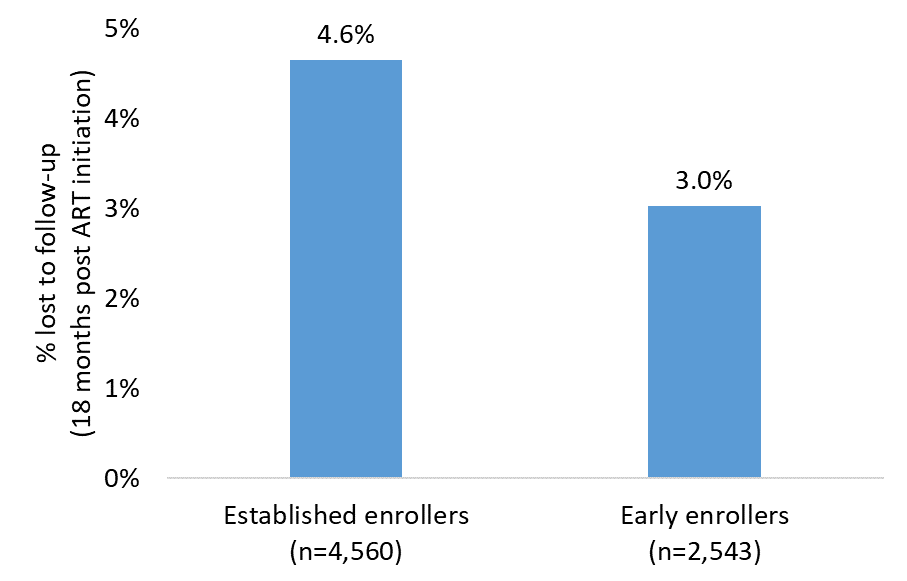

A potential area of concern was that facilities that had better-than-average retention would be more willing or able to enroll patients into DSD models early and therefore skew the results. We therefore conducted this sub-analysis where we limited the data to those facilities which had substantial proportion of their patients enrolled into DSD models early. Criteria for this analysis limited the data to facilities where: i) ≥20% of patients had early enrollers, and ii) at least 100 patients across both groups (early enrollers and established enrollers). 37 facilities across 8 of 10 provinces were selected for this analysis; 73% (n=27) of facilities were in urban areas. This analysis consisted of 7,103 patients: majority (61%, n=4,351) were female, age group distribution was similar to the main analysis (Table 2) (11%, n=784 were 15-24 years; 35%, n=2,488 were 25-34 years; 43%, n=3,028 were 35-49 years; 11%, n=799 were 50+ years), 81% (n=5,731) of patients were in urban settings. Majority (57%, n=4,028) of patients were enrolled into multi-month dispensing, 29% (n=2,058) were in fast-track, 7% (n=484) were in community pick-up points, 5% (n=350) were in home ART delivery, and <2% were in adherence groups (n=112) and extended clinic hours’ groups (n=71).

Results show that in this subset of clinics, early enrollers were less likely to be lost to follow-up (3.0% [77/2,543]), compared to established enrollers (4.6% [212/4,560]). A log-binomial regression assessing risk of loss to follow-up, adjusting for age, sex, urban/rural status, and ART dispensing period estimated that, compared to established enrollers, early enrollers were 40% less likely to be lost to follow-up; adjusted risk ratios (aRR) 0.60 (95% CI 0.46-0.78).
